# The effectiveness of Evodia rutaecarpa hot compress on the recovery of gastrointestinal function after laparoscopic surgery for colorectal cancer: A propensity score-matched retrospective cohort study

**DOI:** 10.1101/2024.05.07.24306986

**Authors:** MiaoXin Huang, JunMiao Li, Wei Huang, YuLin Zhou, Lei Cai, Ming Liu

**Author notes:** Peking University Health Science Center - Macao Polytechnic University Nursing Academy, Macao Polytechnic University, Macao SAR, China. The Second Affiliated Hospital of Guangzhou University of Chinese Medicine, Guangzhou, Guangdong Province, China. These authors contributed equally to this work (MiaoXin Huang, JunMiao Li). These authors also contributed equally to this work (Wei Huang, YuLin Zhou).

## Abstract

**Background:** Although the use of hot compresses with the herbal medicine Evodia rutaecarpa (ER) as a complementary and alternative therapy to promote recovery of postoperative gastrointestinal function is gradually increasing in clinical practice, there is still a lack of relevant empirical studies. Particularly, the role of ER hot compress therapy on gastrointestinal recovery post-laparoscopic surgery for colorectal cancer has not been well investigated. The purpose of this study is to evaluate the efficacy and applicability of ER hot compress therapy for the recovery of postoperative gastrointestinal function.

**Methods:** This is a retrospective cohort study. Patients were divided into two cohorts, the ER group and the non-ER group. Propensity score matching(PSM) was introduced to limit confounding, and independent samples t-tests, non-parametric tests, or Chi-squared tests were used to compare these two cohorts.

**Results:** A total of 454 patients were included, with 267 (59%) receiving ER hot compress therapy and 187 (41%) not. Following 1:1 PSM, 320 patients were analyzed (160 in each group). Compared to the ER group, patients in the non-ER group had shorter times to return to a semi-liquid diet (p=0.035) and hospital stay (p=0.001), as well as lower hospital costs (p<0.001). Subgroup analyses revealed no statistically significant differences in the length of hospital stay, hospital costs, postoperative time to return to full-liquid diet, or time to return to semi-liquid diet among stage I and II tumor patients, though means and standard deviations were generally lower in the ER group. Complication incidence showed no significant difference between the two cohorts before and after PSM.

**Conclusions:** The use of ER hot packs after laparoscopic surgery in patients with colorectal cancer has a non-significant effect on the recovery of the gastrointestinal function and, given the results of the study, it is likely that patients with early-stage tumors may benefit more. Therefore, healthcare providers need to consider the individualisation, practicality, and economics of treatment options.

## Introduction

The World Health Organization (WHO) has released the latest estimates of the global cancer burden, showing that colorectal cancer (CRC) is the third most common cancer worldwide (over 1.9 million cases, 9.6%) and the second leading cause of cancer deaths (over 0.9 million cases, 9.3%)[1]. In addition, the World Health Organization’s colorectal cancer epidemiological data for the whole of 2022 show that Asia ranks first in terms of colorectal cancer incidence, mortality, and 5-year prevalence (colon, rectum), followed by Europe with the second highest share[2]. Studies have shown that the incidence and mortality of CRC have decreased in some European, Oceanian and North American countries in recent years [3–5], but data from the Chinese Cancer Center in 2022 show that the incidence and mortality of CRC continue to increase [3, 6–7]. Worryingly, the incidence of colorectal cancer is steadily increasing in China and globally in people under the age of 50 [8–14]. Therefore, the prevention and treatment of colorectal cancer are urgent, and various treatment regimes and technologies have emerged. The surgical procedures have been moved from traditional open surgery to laparoscopic surgery and robotic-assisted laparoscopic surgery.

However, postoperative colorectal cancer patients still face various challenges. Among them, postoperative gastrointestinal dysfunction (POGD) is one of the common complications of abdominal surgery, including laparoscopic colorectal cancer surgery [15]. The most commonly occurring symptoms of POGD are nausea, vomiting, abdominal distention, postoperative intestinal ileus (POI), gastrointestinal bleeding, and possibly progressive multi-organ failure[5]. Therefore, a variety of complementary and alternative therapies have been used to help restore gastrointestinal function after surgery, particularly a range of herbal medicines.

In the past few years, Evodia rutaecarpa (ER), also known as Wuzhuyu (a type of herbal medicine), has received increasing attention from various health professionals in mitigating discomfort and supporting recovery from POGD. ER was first described more than 2000 years ago in the Shennong Ben Cao Jing (a Chinese book on agriculture and medicinal plants)[16]. Nowadays, herbal therapy has attracted worldwide attention[17]. As an herbal remedy with potential value for CRC patients, ER is worth exploring in depth. In China, the use of ER hot compress therapy is becoming increasingly popular..

Based on the literature review, a study by Li and his colleagues showed that the evodiamine and the rutaecarpine derived from ER have high permeability and remarkable selective transdermal properties[18]. Furthermore, Gu and his colleagues stated that sesquiterpenoids have significant pharmacological activities and suggested that the volatile oil of Cornus officinale can pass through PTGS1, PTGS2 and IL-6[19]. Because of these properties, ER is mixed with coarse salt in equal proportions to make an ER hot compress bag in clinical practice, heated in a microwave oven at medium heat for 4 minutes, and then applied to the patient’s navel or Shenque point.

In the implementation of complementary and alternative therapies, not only doctors and rehabilitation therapists, but also nurses play an essential role. Therefore, when implementing complementary and alternative therapies, nurses should proactively understand their efficacy and applicability, use scientific and effective nursing practice based on evidence-based nursing, and strive to be patient advocates.

Therefore, this retrospective cohort study focused on colorectal cancer patients undergoing laparoscopy to investigate the effect of ER hot compress therapy on the recovery of postoperative gastrointestinal function.

## Materials and method

### Study design and data source

This was a retrospective cohort study based on the Strengthening the Reporting of Observational Studies in Epidemiology (STROBE) guidelines (See S1 File.)[20–21]. The study was approved by the Ethics Committee of Macao Polytechnic University (approval number: FCSD/MSN-0051/2023) and the Institutional Review Board of the Second Affiliated Hospital of Guangzhou University of Chinese Medicine (approval number: ZE2023-459-01), and individual written informed consent was not required. The study was conducted in accordance with the principles of the Declaration of Helsinki, and the investigators collected electronic medical records of colorectal cancer patients who underwent laparoscopic surgery at the Second Affiliated Hospital of Guangzhou University of Chinese Medicine (Guangdong Provincial Hospital of Chinese Medicine) from December 1, 2016 to December 30, 2022, including patient demographics, perioperative data, and other information. Throughout the study, patient data were de-identified (recorded and stored in a coded form).

### Study sample

All data was collected from December 21st, 2023 to February 10th, 2024. The study population was selected based on the following inclusion criteria: 1) meet the diagnostic criteria for CRC in the 2017 edition of the Chinese Standards for the Diagnosis and Treatment of CRC and further confirmed by pathological biopsy[22]; 2) all participants underwent elective standard laparoscopic radical tumor curettage; 3) the age of patients was between 25 and 80 years; 4) participants signed informed consent for surgery and use of complementary and alternative therapies (herbal ER hot iron therapy).

The exclusion criteria for the study were: 1) a confirmed diagnosis of distant metastasis of the tumor; 2) combination with other malignant tumors; 3) physical disability that prevents cooperation with functional exercises; 4) mental illness or cognitive impairment that makes communication with the participant difficult; 5) pregnancy or breastfeeding; 6) patients undergoing abdominal surgery within 6 months and having severe intestinal adhesions; 7) patients with unsatisfactory control of the primary medical disease; 8) concurrent other serious organic diseases; 9) preoperative abdominal infection; 10) patients requiring postoperative blood transfusion; 11) patients requiring open surgery or extended radical surgery; 12) undergoing combined abdominal perineal radical rectal cancer surgery (Miles procedure); 13) patients experiencing serious complications within 6 hours of surgery or requiring admission to intensive care after surgery; 14) patients with allergy to herbal ER therapy.

### Definition and measurement of exposure factors and outcome indicators

Patients were divided into two groups according to the exposure factor (i.e. whether they received ER hot compress treatment after laparoscopic surgery for CRC): the ER group and the conventional treatment group (the non-ER group).

1. The non-ER group only received routine postoperative care, including monitoring of vital signs, postoperative fluid restoration, care of drainage tubes and monitoring of drainage fluids, pain relief, surgical wound care and daily care.
2. The ER group received ER hot compress therapy and routine postoperative care. 250 grams of ER (from Guangdong Kangmei Pharmaceutical Co., Ltd.) and 250 grams of coarse salt (Guangzhou Salt Industry Co., Ltd.) were placed in a cotton bag of 18 cm x23 cm and heated in a microwave oven at medium heat for 2 to 3 minutes. The temperature of the ER hot pack was about 60 - 70 ℃, and tolerable to the patient. Nurses massage the ER hot pack in a clockwise direction with the Shenque point (navel) as the centre for 20 circles, avoiding the surgical incision. The ER hot compress treatment is usually started 24 hours after surgery, twice a day for 20 minutes each time.
3. Postoperative gastrointestinal outcomes and their measurements: time of first postoperative flatus, time of first postoperative defecation, and time of recovery of postoperative bowel sounds (the actual measurement of the above three items is recorded from the end of the operation to the time when the doctor asks the patient about the event, and it is not possible to calculate the exact time when the patient has the above events), time of full-liquid diet and time of semi-liquid diet (the actual measurement time is from the end of the operation to the time when the doctor gives the order, i.e. the time when the patient starts the full-liquid diet) and the incidence of postoperative gastrointestinal complications (including postoperative nausea and vomiting, abdominal pain and bloating, anastomotic fistula, and bowel paralysis and obstruction).

### Covariates

The selection of covariates consists of three steps in this study. First, possible covariates were selected based on the investigator’s clinical experience and literature review, including surgical site, tumor stage, history of chronic gastrointestinal disease, intestinal stoma, gender, chronic lung disease, anaemia, hypoproteinaemia, history of abdominal surgery, diabetes mellitus, hypertension, hyperlipidaemia, history of heart disease, history of smoking, history of alcohol consumption, age, BMI, intraoperative bleeding, and operative time. Second, the raw data were statistically analysed to screen for possible confounders based on the grouping variable (whether ER hot compress therapy was used or not). Third, different caliper values were adjusted in the R language software to analyse which covariates were likely to have an effect on the study results. The main covariates examined were: 1) continuous variables: age and intraoperative bleeding; 2) categorical variables: hyperlipidaemia and tumor stage.

### Handling of missing data

Firstly, the type of data missing from the Electronic Medical Record System(EMR) information was analysed, such as the patient’s basic information, BMI, and the time of the patient’s first post-operative bowel movement; Secondly, the analysis concluded that the reason for the missing data collected in this study was Missing Completely at Random (MCAR), due to human factors that were not recorded, omitted, or lost, and did not affect the unbiased nature of the sample: 1) it was divided into the following cases, and if there were more, the researcher simply deleted them; 2) if there were fewer missing values, those missing values were filled by interpolation using the mean and the plurality of the missing values.

### Statistical methods

R4.2.3 software and SPSS 27.0 were used for statistical analysis. Measures that conformed to normal distribution and homogeneity of variances were expressed as mean standard deviation (mean ± SD) and independent t-test was used for comparison between two groups, while those that did not conform to normal distribution were expressed as median (interquartile range), i.e. M (Q1, Q3) and Mann-Whitney U-test was used for comparison between two groups. Count data were expressed as cases (%) and comparisons between two groups were made using the Pearson chi-squared test or Fisher’s exact test. All statistical inferences were made using two-tailed tests with a statistically significant test level of ɑ=0.05 and 95% CI for parameter CI estimation.

### Propensity score matching

To assess the impact of confounding bias due to non-randomisation in observational studies, propensity score matching (PSM) was proposed in this study[23–24]. Four variables, tumor stage, hyperlipidaemia, intraoperative bleeding and age, were used as covariates, and the use of ER hot packs was used as a grouping variable(exposure factor). Propensity scores were calculated for each sample using nearest neighbour 1:1 matching and a caliper value of 0.05. In addition, the investigators visually compared the distribution difference of the two data sets before and after matching by plotting distribution plots, density plots, and empirical cumulative distribution function (eCDF) plots, respectively.

### Sensitivity analysis

First, the parameters of the sensitivity analysis were determined, and common parameters included caliper values and matching ratios[25]. The researcher evaluates and compares the matching results under different caliper values (e.g. selecting different caliper values of 0.02, 0.03, 0.05, 0.1, and 0.2; as well as selecting different matching ratios (e.g. ratio=1 and ratio=2) and also compares the matching results under different matching variables).

Second, the robustness and sensitivity of the PSM results can be assessed by comparing the sample sizes after matching under different parameter settings by statistically analysing the results, and by assessing the distribution of the scores after matching by plotting distribution plots and histograms. It is assumed that the variation of the matching results under different parameter settings and different matching variables is small, which indicates that the matching results are less sensitive to the choice of parameters and have better robustness and sensitivity.

### Equilibrium test

Equilibrium tests are used to assess whether the baseline characteristics between the two data groups (ER and non-ER groups) before and after PSM are effectively balanced. Commonly used equilibrium tests include the Standardised Mean Difference (SMD), the Chi-square test, and the t-test[26–27]. The SMD was used in this study to assess the effect of adjustment. If the SMD value is small (less than 0.1), it means that the matching effect is good, the confounding factors between the two groups have been effectively controlled, and the research results are credible. In addition, SMD plots and histograms were used to visually analyse the baseline distribution of data between the two groups before and after matching.

## Subgroup analysis

To further explore the heterogeneity of the samples after PSM, to analyse the variability of different treatments, and to improve the accuracy and reliability of the results, the researchers performed subgroup analyses of the pooled data. Because tumor staging is usually clinically relevant to the patient’s postoperative treatment plan. Therefore, according to the American Joint Committee on Cancer (AJCC) cancer staging system[28] and the diagnosis of clinicians, tumor staging was divided into different subgroups for analysis.

## Results

### The process of study population selection

Before matching, there were 267 cases in the ER group and 187 cases in the non-ER group. Statistical analysis was performed on the final 454 patients enrolled (statistical values represent T value, U value, and chi-square value, respectively) . This analysis indicated that, firstly, the p-values for age and hyperlipidemia were 0.015 and 0.008, respectively, indicating statistical significance and potential impact on the results. Secondly, compared to the non-ER group, patients in the ER group had higher hospital costs (p < 0.001), longer hospital stays (p < 0.001), and longer time to resume a semi-liquid diet after surgery (p = 0.021). The remaining results showed no statistical significance (see Table 1 and S.Table 1) and other additional information is provided in S.Table 2.

**Table 1.**
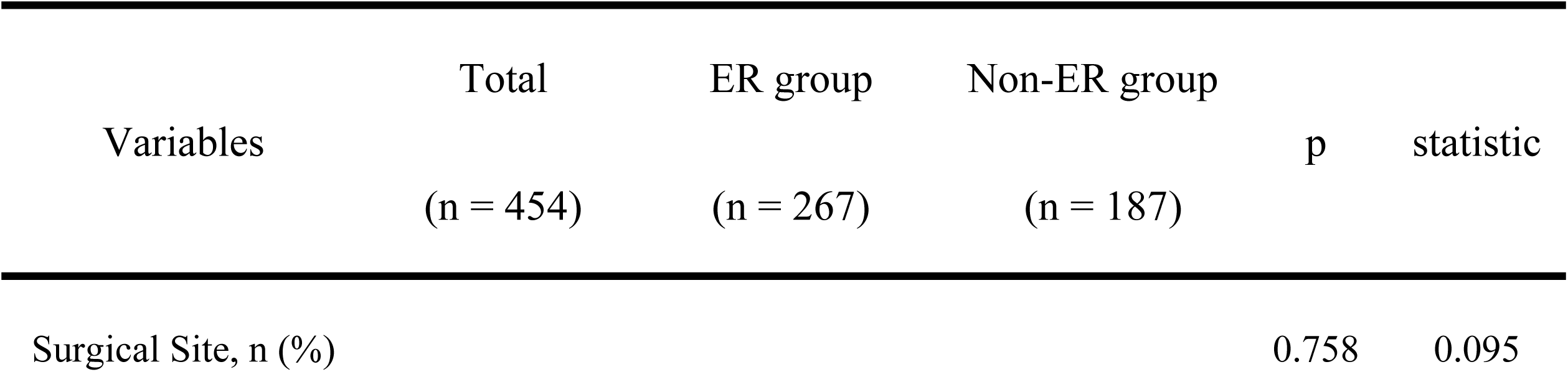
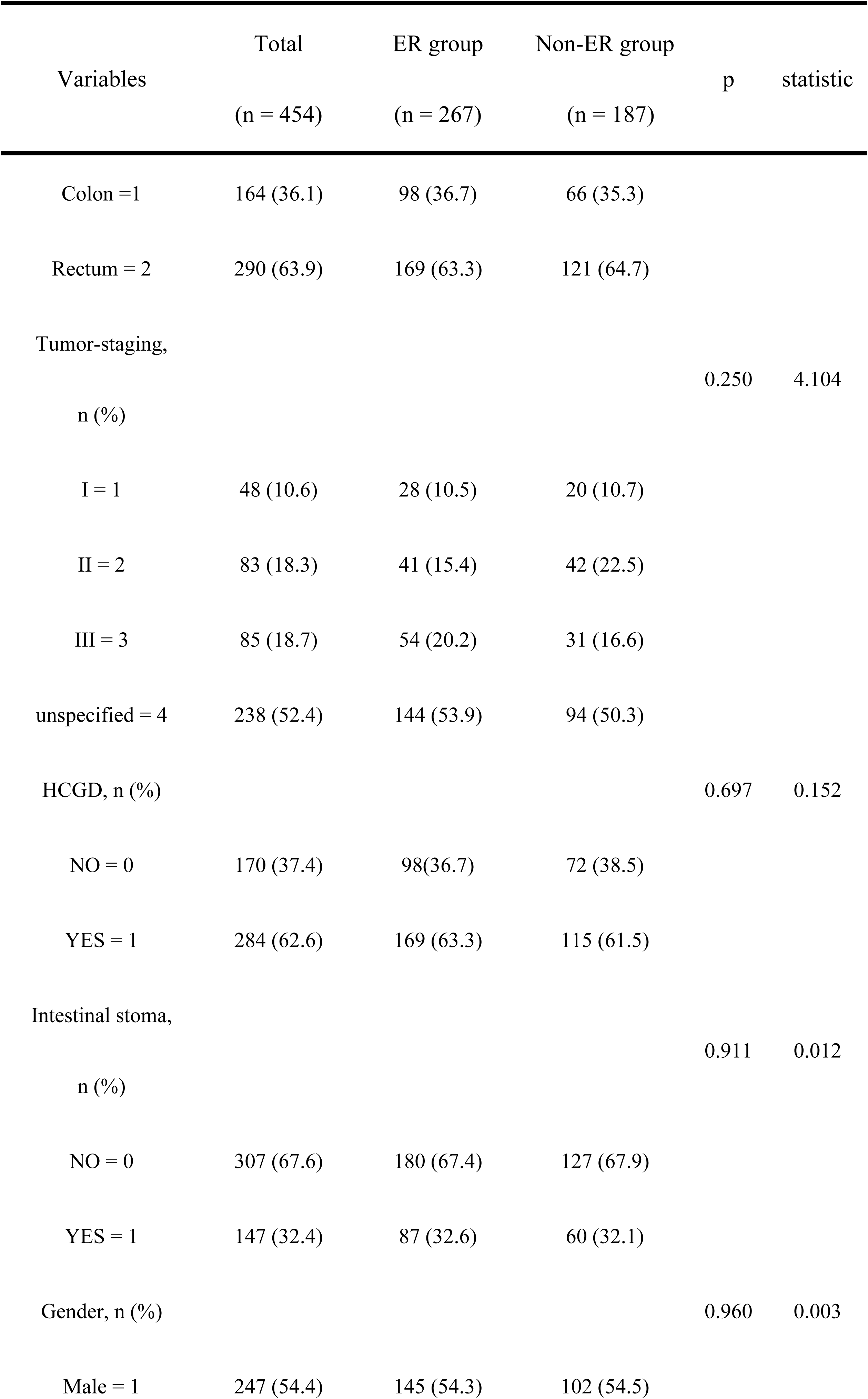
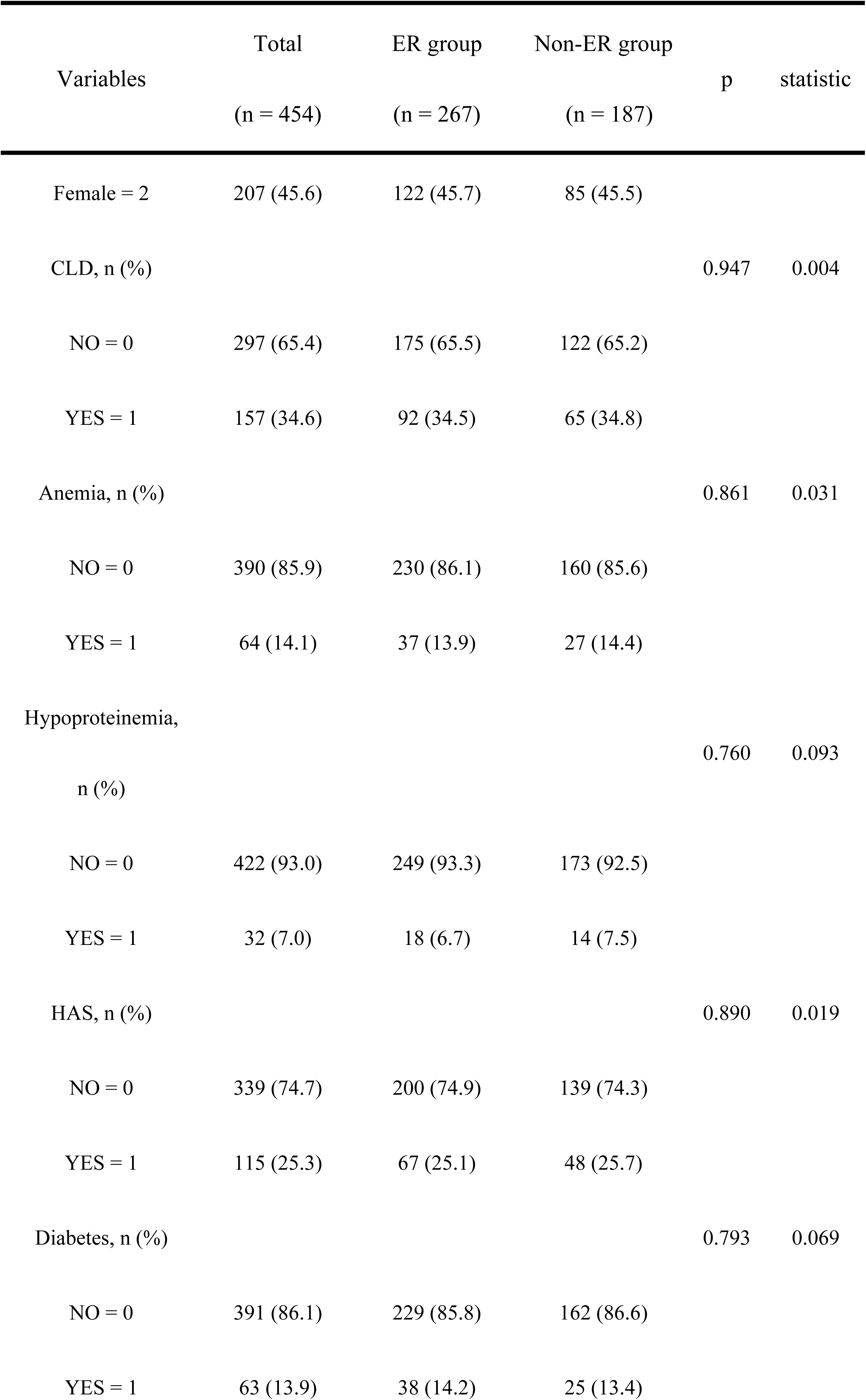
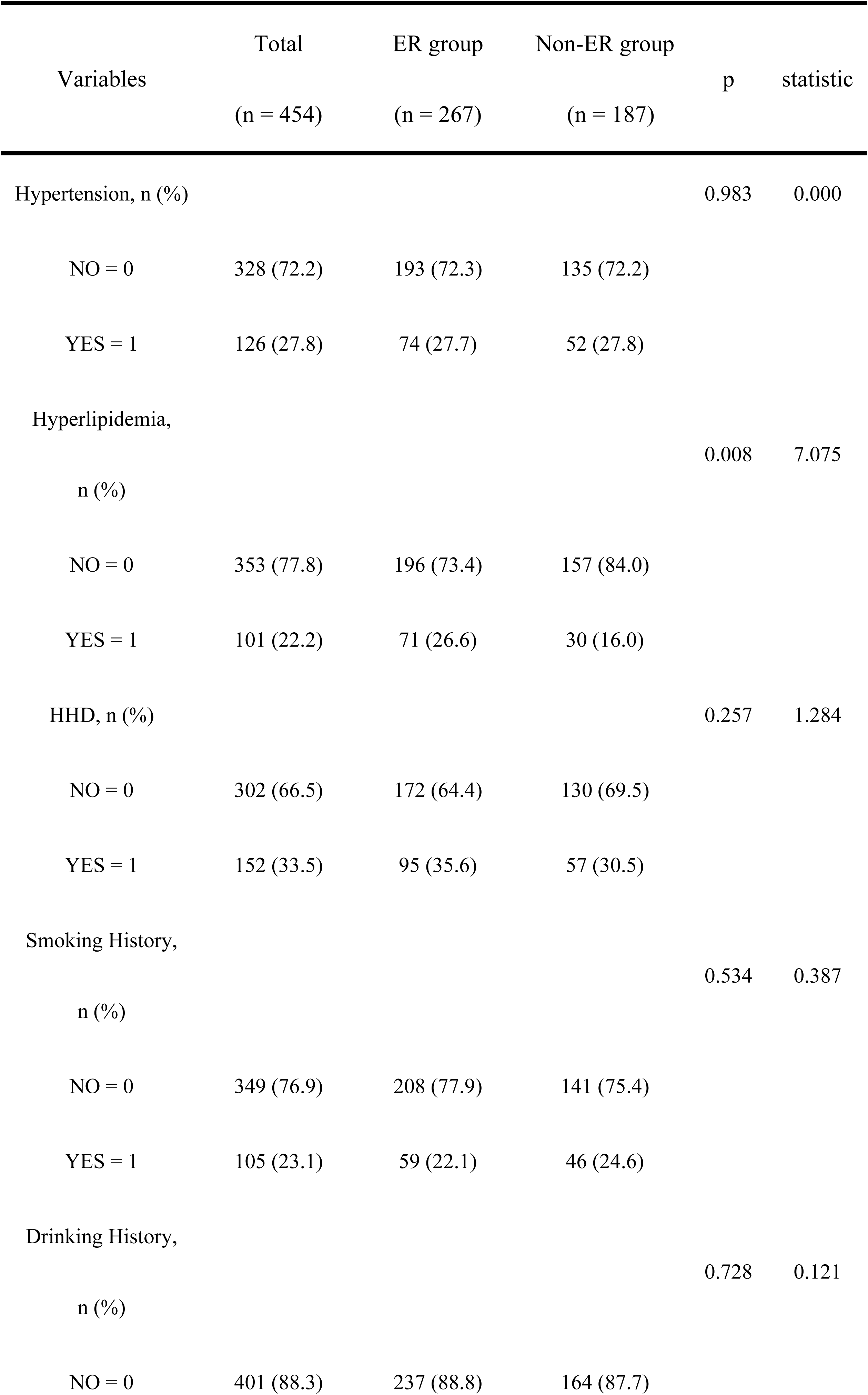
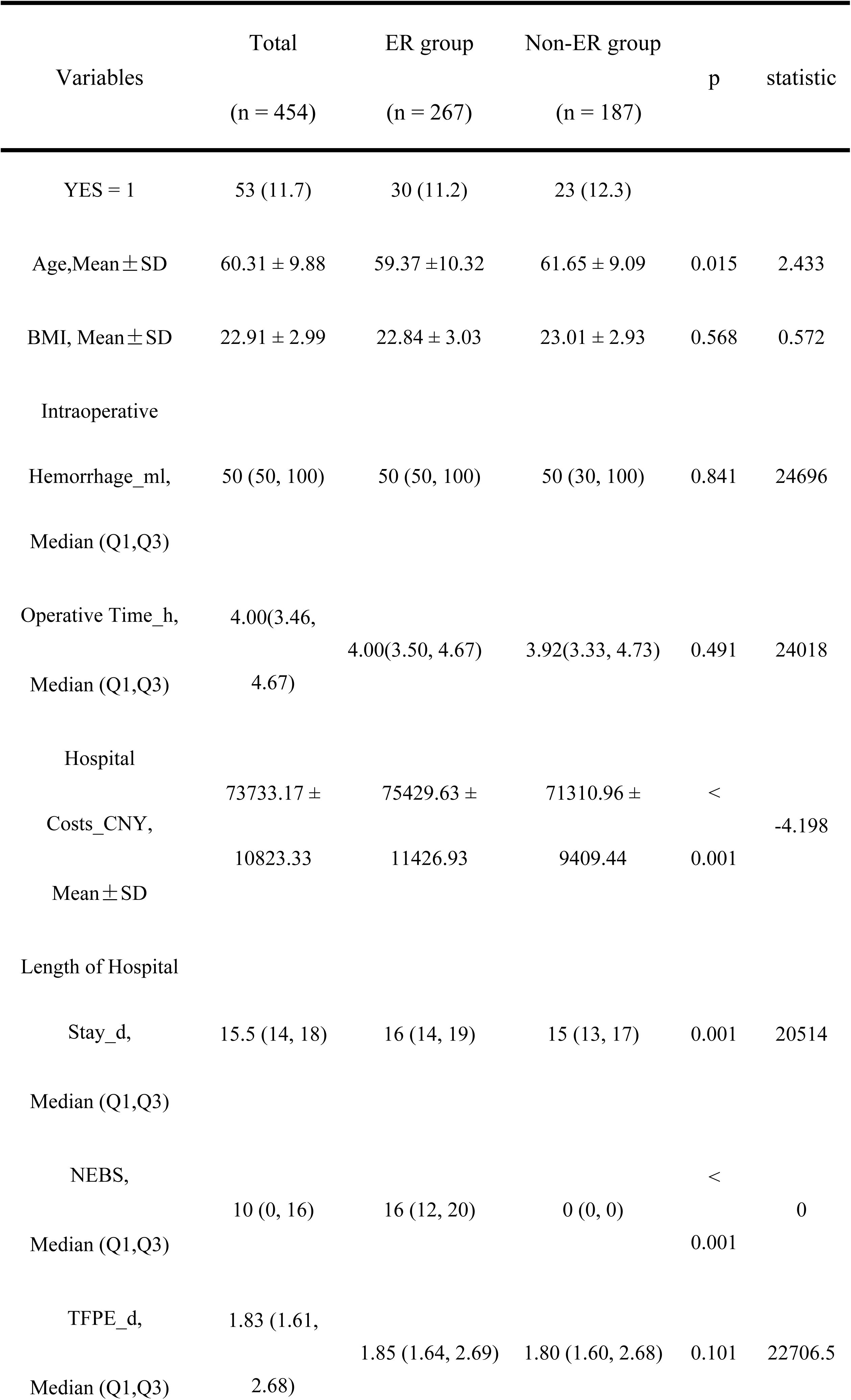
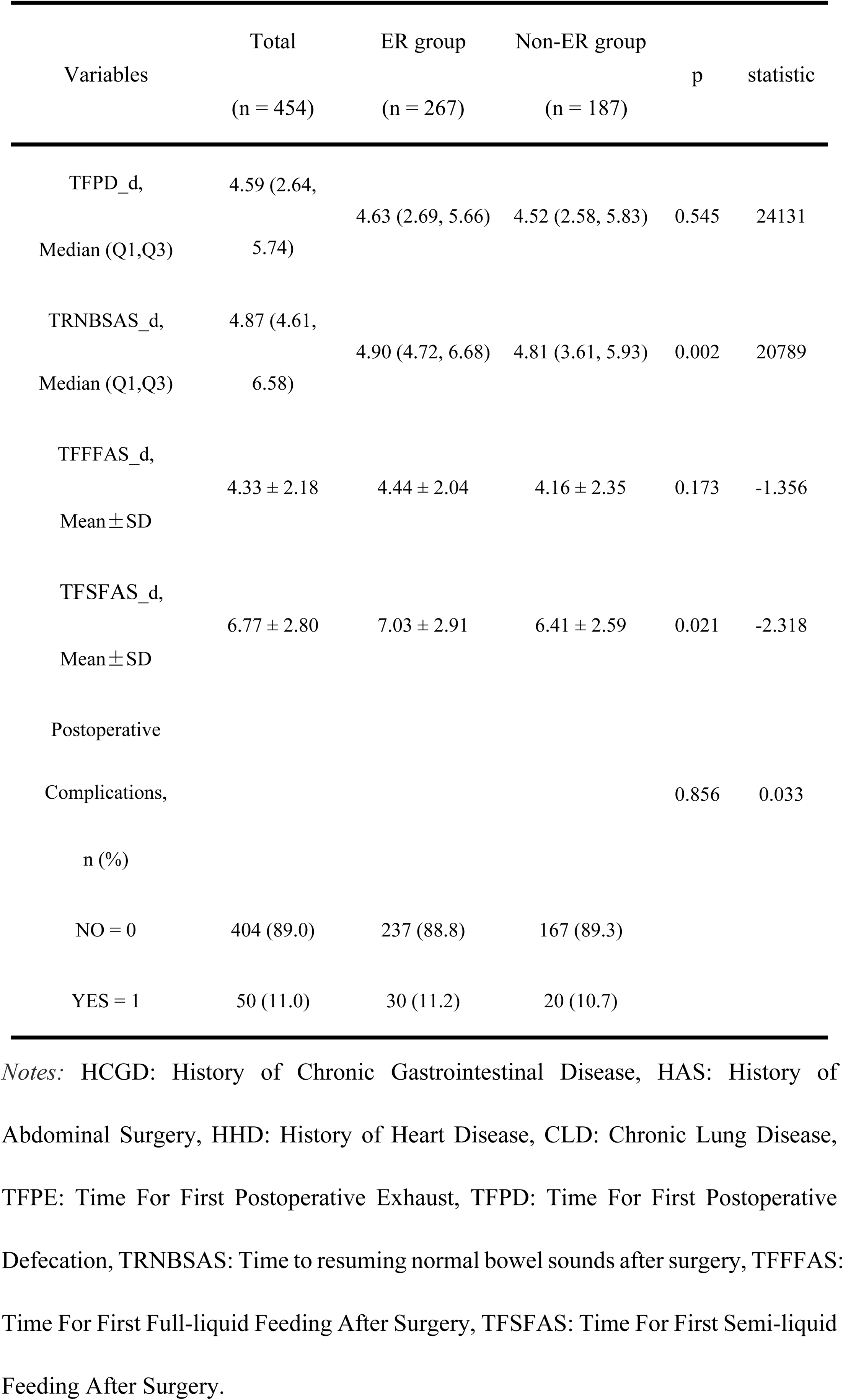
Baseline data before PSM.

### After PSM

The data of 454 patients underwent propensity score matching (PSM) analysis using the nearest neighbor method, with age, tumor stage, intraoperative hemorrhage, and hyperlipidemia as matching variables. Age and hyperlipidemia were identified as potential confounding factors before matching, while tumor stage and intraoperative hemorrhage became statistically significant after adjusting for a smaller caliper value and were thus included as matching variables. The matching ratio was 1:1, with a caliper set at 0.05, resulting in successful matching of 320 patients, including 160 in the ER group and 160 in the non-ER group.

Before PSM, there were significant differences between the ER and non-ER groups in terms of age and hyperlipidemia (with p-values of 0.041 and 0.008, respectively), indicating potential confounding factors. However, after PSM, the differences between the ER and non-ER groups in all characteristics became non-significant (all P>0.05, as detailed in Table 2), indicating successful achievement of balanced comparability after PSM.

**Table 2.** Baseline data after PSM.

| Variables | Total<br>(n = 320) | ER group<br>(n = 160) | Non-ER<br>group<br>(n = 160) | p | statistic |
| --- | --- | --- | --- | --- | --- |
| Surgical Site, n (%) |  |  |  | 0.818 | 0.053 |
| Colon =1 | 122 (38.1) | 62 (38.8) | 60 (37.5) |  |  |
| Rectum = 2 | 198 (61.9) | 98 (61.3) | 100 (62.5) |  |  |
| Tumor staging, n (%) |  |  |  | 1.127 | 0.771 |
| I = 1 | 30 (9.4) | 13 (8.1) | 17 (10.6) |  |  |
| II = 2 | 57 (17.8) | 30 (18.8) | 27 (16.9) |  |  |
| III = 3 | 54 (16.9) | 25 (15.6) | 29 (18.1) |  |  |
| unspecified = 4 | 179 (55.9) | 92 (57.5) | 87 (54.4) |  |  |
| HCGD, n (%) |  |  |  | 0.251 | 1.317 |
| NO = 0 | 124 (38.8) | 57 (35.6) | 67 (41.9) |  |  |
| YES = 1 | 196 (61.3) | 103 (64.4) | 93 (58.1) |  |  |
| Intestinal Stoma,<br>n (%) |  |  |  | 0.545 | 0.366 |
| NO = 0 | 221 (69.1) | 113 (70.6) | 108 (67.5) |  |  |
| YES = 1 | 99 (30.9) | 47 (29.4) | 52 (32.5) |  |  |
| Gender, n (%) |  |  |  | 0.575 | 0.314 |
| Male = 1 | 171 (53.4) | 88 (55.0) | 83 (51.9) |  |  |
| Female = 2 | 149 (46.6) | 72 (45.0) | 77 (48.1) |  |  |
| CLD, n (%) |  |  |  | 1.000 | 0 |
| NO = 0 | 210 (65.6) | 105 (65.6) | 105 (65.6) |  |  |
| YES = 1 | 110 (34.4) | 55 (34.4) | 55 (34.4) |  |  |
| Anemia, n (%) |  |  |  | 0.623 | 0.242 |
| NO = 0 | 277 (86.6) | 140 (87.5) | 137 (85.6) |  |  |
| YES = 1 | 43 (13.4) | 20 (12.5) | 23 (14.4) |  |  |
| Hypoproteinemia, |  |  |  | 1.000 | 0 |
| n (%) |  |  |  |  |  |
| NO = 0 | 302 (94.4) | 151 (94.4) | 151 (94.4) |  |  |
| YES = 1 | 18 (5.6) | 9 (5.6) | 9 (5.6) |  |  |
| HAS, n (%) |  |  |  | 0.899 | 0.016 |
| NO = 0 | 237 (74.1) | 118 (73.75) | 119 (74.4) |  |  |
| YES = 1 | 83 (25.9) | 42 (26.25) | 41 (25.6) |  |  |
| Diabetes, n (%) |  |  |  | 0.741 | 0.110 |
| NO = 0 | 278 (86.9) | 140 (87.5) | 138 (86.3) |  |  |
| YES = 1 | 42 (13.1) | 20 (12.5) | 22 (13.8) |  |  |
| Hypertension, n (%) |  |  |  | 0.899 | 0.016 |
| NO = 0 | 237 (74.1) | 118 (73.75) | 119 (74.4) |  |  |
| YES = 1 | 83 (25.9) | 42 (26.25) | 41 (25.6) |  |  |
| Hyperlipidemia,<br>n (%) |  |  |  | 1.000 | 0 |
| NO = 0 | 260 (81.25) | 130 (81.25) | 130 (81.25) |  |  |
| YES = 1 | 60 (18.75) | 30 (18.75) | 30 (18.75) |  |  |
| HHD, n (%) |  |  |  | 0.228 | 1.455 |
| NO = 0 | 220 (68.75) | 105 (65.6) | 115 (71.9) |  |  |
| YES = 1 | 100 (31.25) | 52 (34.4) | 45 (28.1) |  |  |
| Smoking History,<br>n (%) |  |  |  | 0.789 | 0.072 |
| NO = 0 | 248 (77.5) | 125 (78.1) | 123 (76.9) |  |  |
| YES = 1 | 72 (22.5) | 35 (21.9) | 37 (23.1) |  |  |
| Drinking History,<br>n (%) |  |  |  | 0.210 | 1.572 |
| NO = 0 | 285 (89.1) | 146 (91.3) | 139 (86.9) |  |  |
| YES = 1 | 35 (10.9) | 14 (8.8) | 21 (13.1) |  |  |
| Postoperative<br>Complications, n (%) |  |  |  | 1.000 | 0 |
| NO = 0 | 296 (92.5) | 148 (92.5) | 148 (92.5) |  |  |
| YES = 1 | 24 (7.5) | 12 (7.5) | 12 (7.5) |  |  |
| Age, Mean ± SD | 59.99 ± 9.09 | 59.64 ± 9.43 | 60.34 ± 8.76 | 0.488 | 0.694 |
| BMI, Mean ± SD | 22.89 ± 2.98 | 22.70 ± 3.05 | 23.08 ± 2.90 | 0.259 | 1.132 |
| Intraoperative |  |  |  |  |  |
| Hemorrhage_ml,<br>Median (Q1,Q3) | 50 (50, 100) | 50 (50, 100) | 50 (30, 100) | 0.161 | 11669.5 |
| Operative Time_h,<br>Median (Q1,Q3) | 3.91 (3.42, 4.62) | 3.92(3.50, 4.65) | 3.90 (3.30, 4.61) | 0.507 | 12251.5 |
| Hospital Costs_CNY,<br>Mean±SD | 72435.17 ± 10123.06 | 74856.62 ± 11208.38 | 70013.72 ± 8256.44 | < 0.001 | -4.400 |
| Length of Hospital |  |  |  |  |  |
| Stay_d,<br>Mean±SD | 15.74 ± 3.85 | 16.46 ± 4.048 | 15.01 ± 3.495 | < 0.001 | -3.429 |
| NEBS,<br>Median (Q1,Q3) | 3 (0, 16) | 16 (12, 19.75) | 0 (0, 0) | < 0.001 | 0 |
| TFPE_d,<br>Median (Q1,Q3) | 1.83 (1.60, 2.63) | 1.86 (1.60, 2.65) | 1.80 (1.60, 2.60) | 0.195 | 11728.5 |
| TFPD_d,<br>Median (Q1,Q3) | 4.59 (2.65, 5.73) | 4.64 (2.78, 5.66) | 4.57 (2.63, 5.82) | 0.439 | 12159.5 |
| TRNBSAS_d, | 4.86 (4.57, | 4.92 (4.67, | 4.80 (3.55, | 0.002 | 10219.5 |
| Median (Q1,Q3) | 6.36) | 6.62) | 5.88) |  |  |
| TFFAS_d,<br>Mean ± SD | 4.15 ± 2.05 | 4.33 ± 1.99 | 3.98 ± 2.10 | 0.129 | -1.522 |
| TFSFAS_d,<br>Mean ± SD | 6.51 ± 2.48 | 6.81 ± 2.58 | 6.20 ± 2.35 | 0.030 | -2.185 |

Additionally, the Jitter Plot (see Fig. 2) illustrated that, after matching, the distributions of matching variables between the ER and non-ER groups tended to be consistent, indicating a good matching effect. The Density Plot and Empirical Cumulative Distribution Function Plot (eCDF plot) (refer to S1 Fig. and S2 Fig.) demonstrated that the density curves or cumulative distribution curves of the two groups’ data were close to each other after matching, further validating the effectiveness of the matching method.

**Fig. 1.**
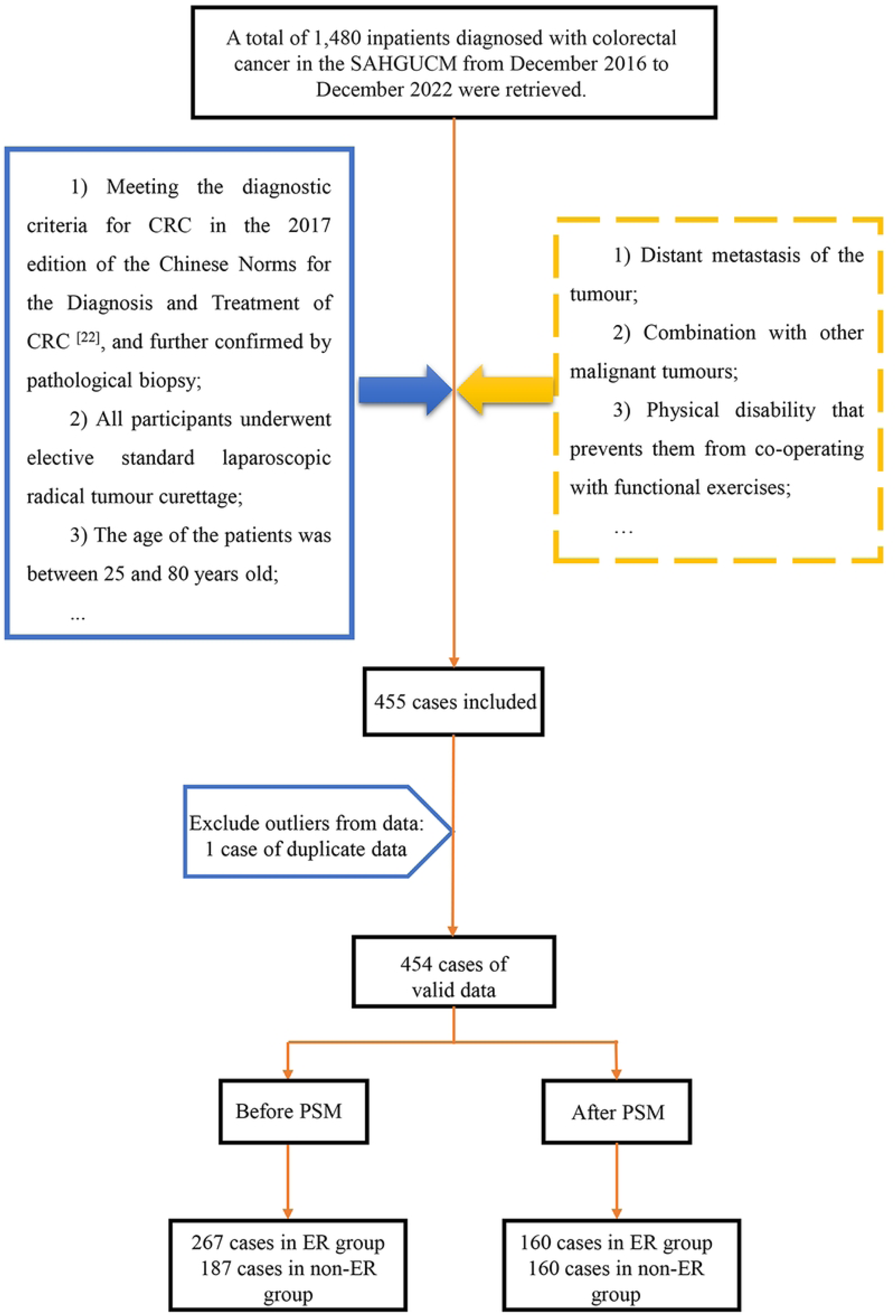
Flowchart of sample selection.

**Fig. 2.**
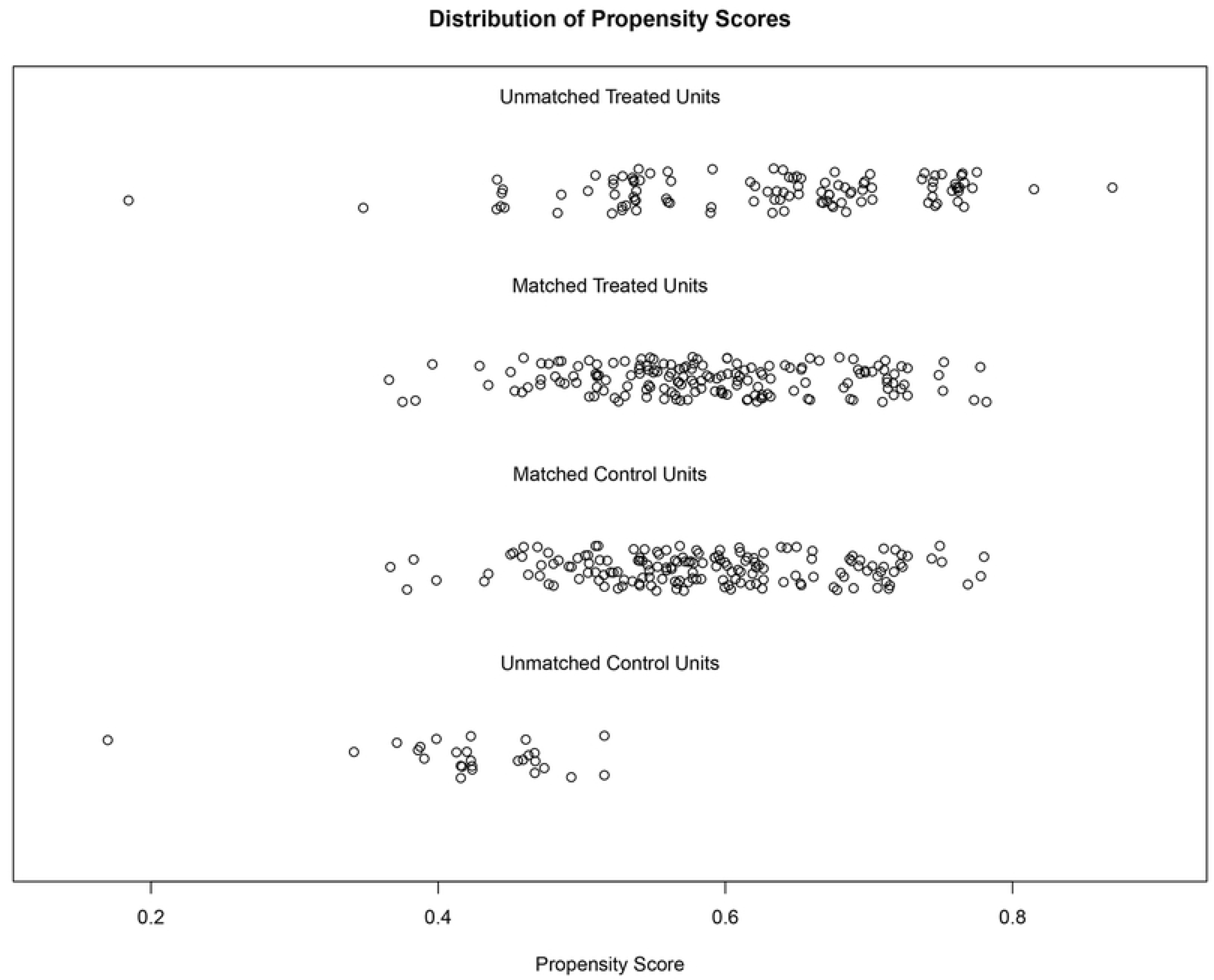
Jitter plot of the ER group and non-ER group after PSM.

As shown in Table 2, after PSM, there were statistically significant differences in postoperative gastrointestinal function recovery indicators between the two groups of patients. Compared to the ER group, the non-ER group had a shorter time to restore bowel sounds postoperatively (p=0.002) and an earlier initiation of a semi-liquid diet (p=0.022). Furthermore, compared to the ER group, the non-ER group had lower hospital costs (p<0.001) and shorter length of stay (p=0.001) (See S1 Table. for additional information).

### Results of sensitivity analysis

The sensitivity analyses revealed that with increasing caliper values and different matching ratio conditions, the sample sizes of the matched ER and non-ER groups exhibited minimal variation. When the ratio was set to 1, as the caliper value increased from 0.02 to 0.2 (caliper values of 0.02, 0.03, 0.1, and 0.2), the sample sizes gradually increased with minor fluctuations, and the cumulative change in the number of cases did not exceed 10% of the total sample size. For instance, the number of matches for the ER and non-ER groups were 139/139, 150/150, 169/169, and 177/177, respectively. Furthermore, when the ratio was set to 2 and the caliper value was 0.05, there were 175 matches for the non-ER group and 160 matches for the ER group. After matching, all covariate p-values were greater than 0.05, indicating consistent results for each match (see S3 Fig. and S4 Fig.).

Hence, this study demonstrates strong robustness in reaching consistent conclusions across various matching conditions, thereby enhancing the reliability and validity of the findings.

### Results of the equilibrium test

The equilibrium and comparability of the two groups of data after PSM was assessed by the Standardised Mean Differences (SMD) before and after matching (see Table 3 and Fig. 3). Before matching, the SMDs of tumor stage, age and hyperlipidaemia exceeded 0.1, indicating the presence of large confounding factors. In contrast, after matching, the SMDs of potential confounders were all less than 0.1, indicating improved sample balance. In addition, the histogram (see Fig. 4) visualised that the histograms of the ER and non-ER groups showed more similar distribution patterns and locations after matching, further indicating that the two groups were more similarly distributed in terms of the characteristic variables and that the PSM was successful in reducing the propensity bias to achieve a balanced result.

**Fig. 3.**
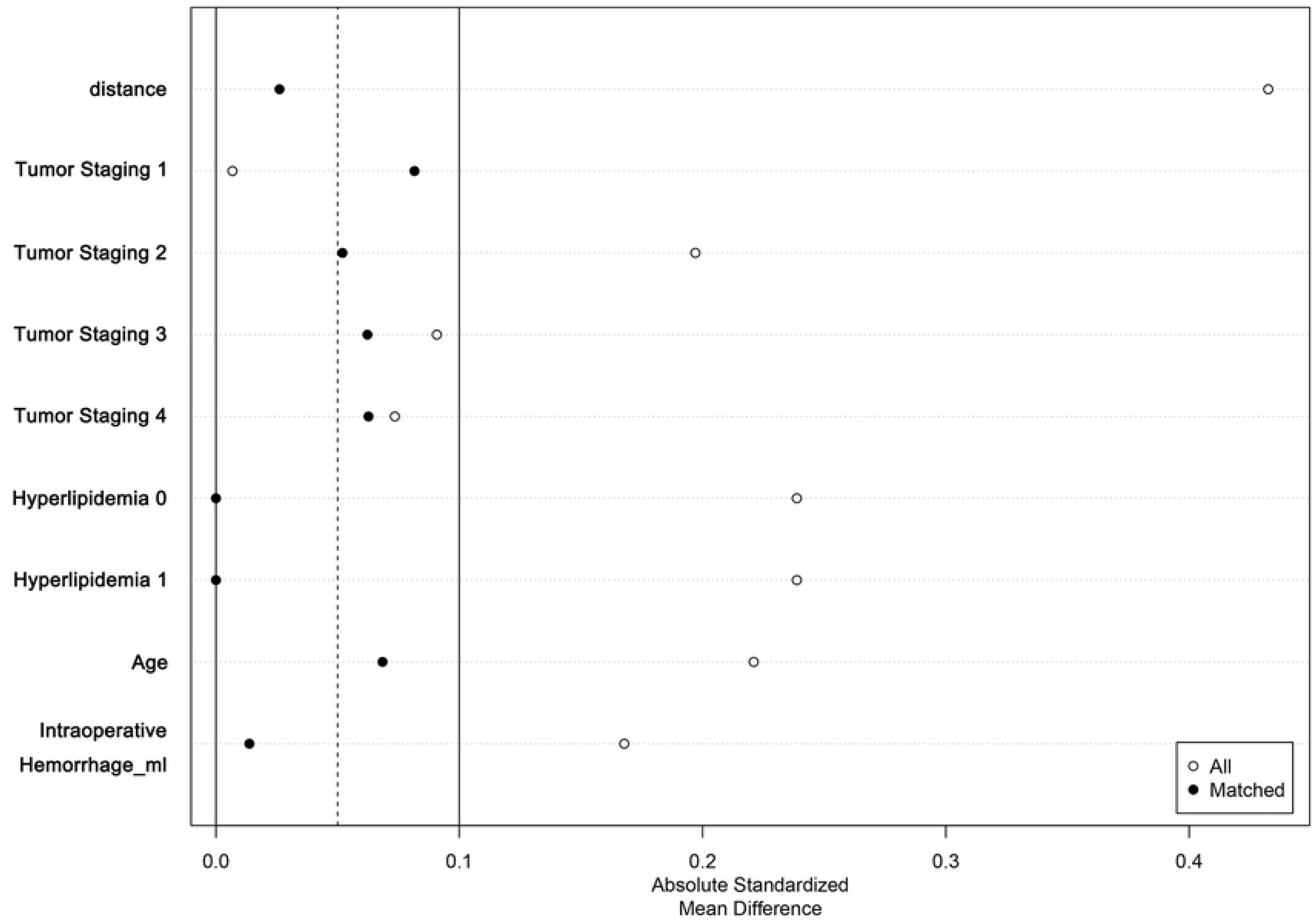
SMD plot of ER group and non-ER group before and after PSM.

**Fig. 4.**
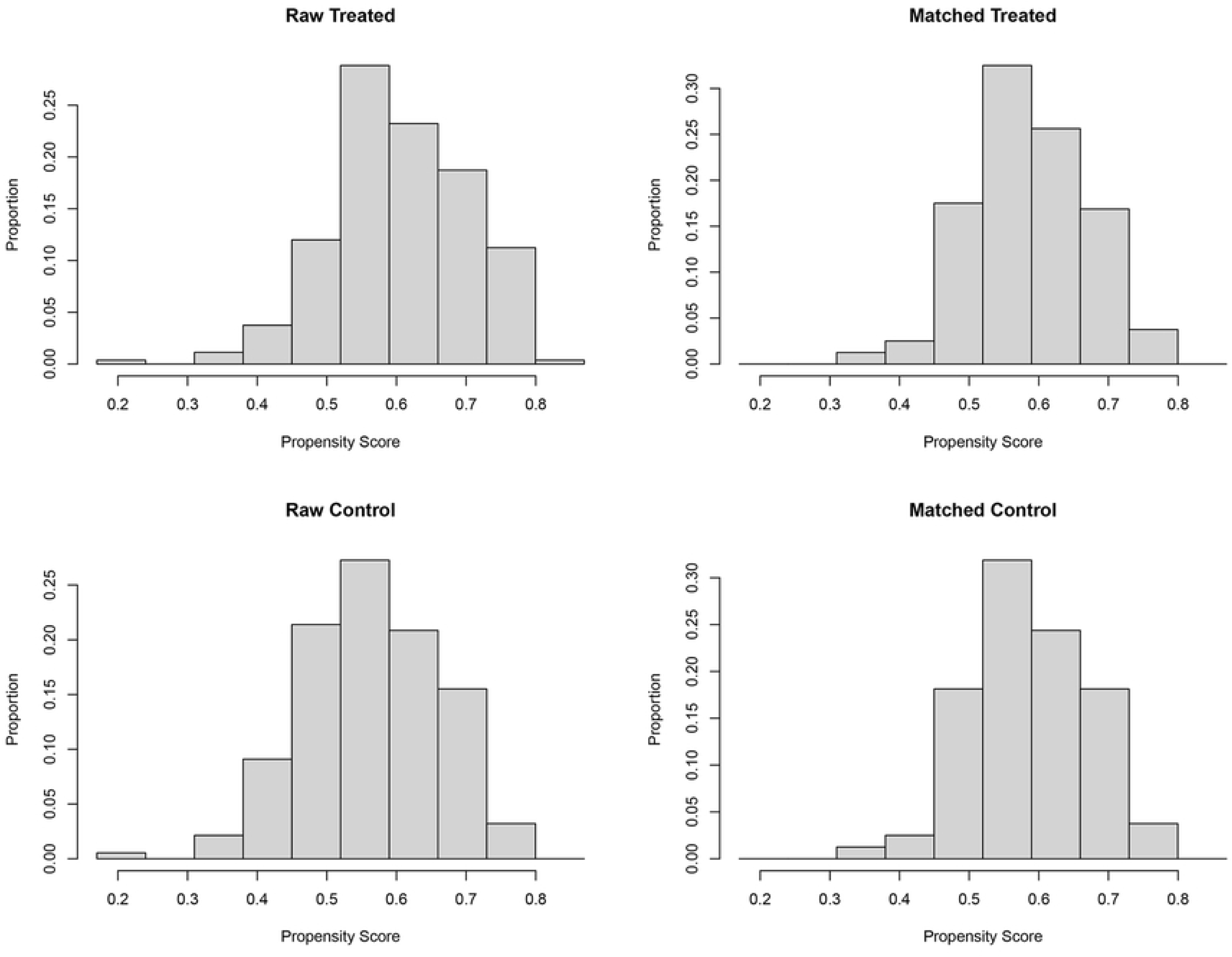
hist graph of ER group and non-ER group before and after PSM.

**Table 3.**
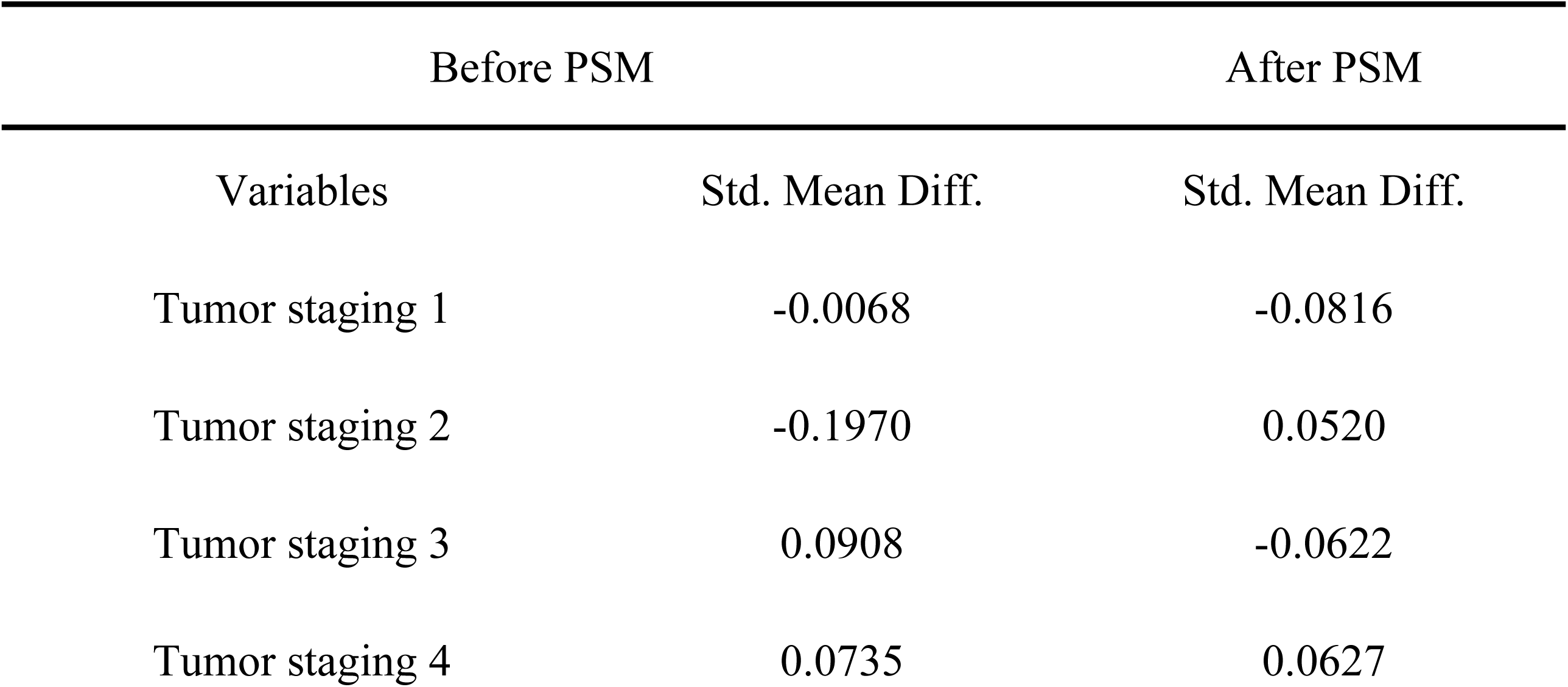
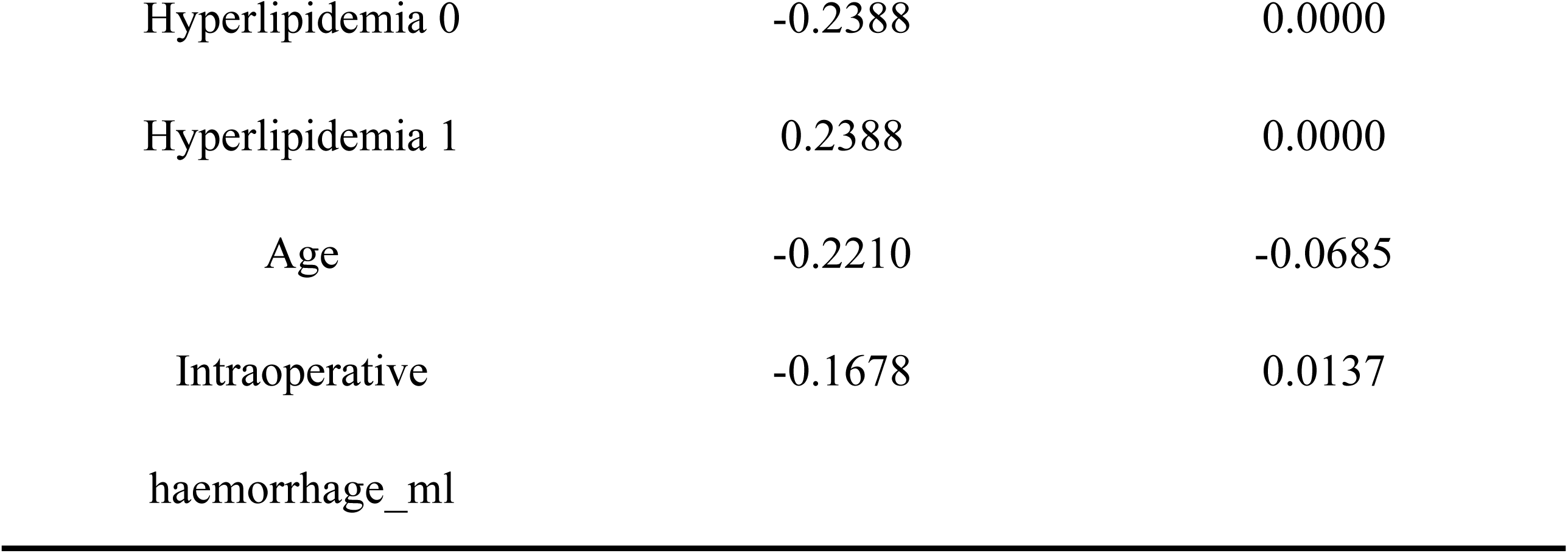
SMD before and after PSM.

### Results of subgroup analysis

This study categorized tumor staging into four subgroups and conducted separate statistical analyses for the ER group and non-ER group.

1. Tumor Stage I: Compared to the non-ER group, patients in the ER group had a slightly longer mean time to full-liquid diet postoperatively (ER: 3.79 ± 1.25 vs non-ER: 3.66 ± 1.20; p=0.765), slightly longer mean time to semi-liquid diet postoperatively (ER: 6.87 ± 2.78 vs. non-ER: 5.70 ± 1.91; p=0.181), slightly longer hospital stay (ER: 16.23 ± 3.30 vs. non-ER: 15.18 ± 2.63; p=0.338), and slightly higher average hospital costs (ER: 75132.22 ± 12657.10 vs. non-ER: 69890.67 ± 11096.58; p=0.238). However, these differences between the groups were not statistically significant (see Table 4-1).
2. Tumor stage II: Except for hospital costs, patients in the ER group had a shorter mean time to full-liquid diet after surgery (ER: 4.15 ± 1.05 vs non-ER: 5.05 ± 3.11; p=0.161), a shorter average time to semi-liquid diet after surgery (ER: 6.40 ± 1.21 vs non-ER: 7.35 ± 3.51; p=0.191), and a shorter average hospital costs (ER: 75576.01 ± 10654.74 vs non-ER: 73310.87 ± 7431.45; p=0.361), and shorter average hospital stay (ER: 16.47 ± 2.69 vs. non-ER: 16.78 ± 4.68; p= 0.756) compared to the non-ER group, although these differences were not statistically significant between groups (see Table 4-2).
3. Tumor stage III and unspecified stage: According to Table 4-3, compared with the ER group, the non-ER group had a significantly shorter postoperative time to semi-liquid diet (ER: 8.35 ± 4.19 vs. non-ER: 6.45 ± 2.33), with a statistically significant difference between the two groups (p=0.041); as shown in Table 4-4, compared with the ER group, the non-ER group not only had a significantly shorter postoperative time to semi-liquid diet (ER: 6.51 ± 2.18 vs. non-ER: 5.87 ± 1.85; p=0.034), but also had significantly lower hospital costs (ER: 75510.44 ± 11584.95 vs. non-ER: 69478.08 ± 7643.31; p<0.001) and shorter hospital stays (ER: 16.29 ± 3.957 vs. non-ER: 14.47± 3.154; p<0.001), with significant statistical differences between the two groups.

**Table 4-1.**
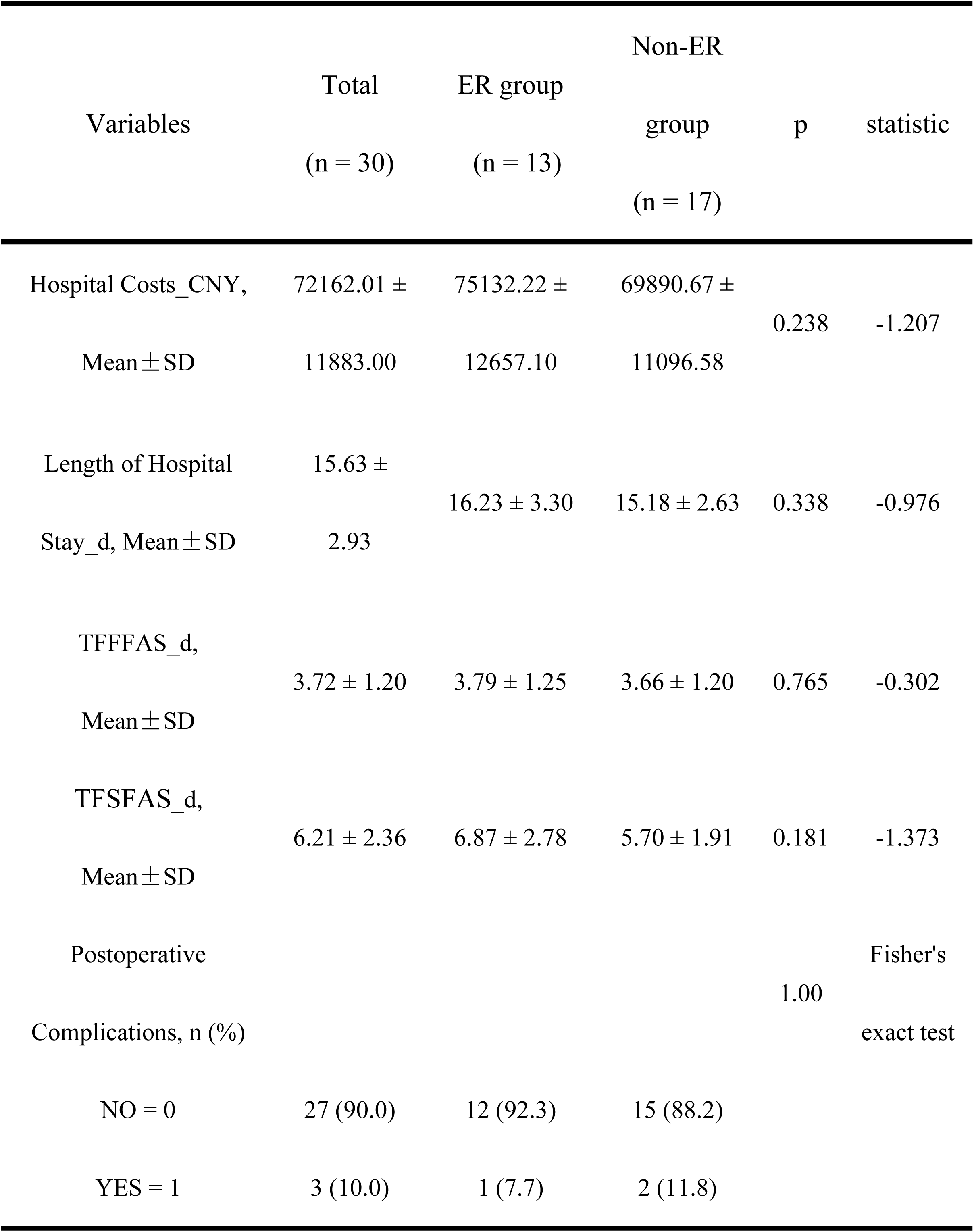
Results of statistical analysis of dependent variables for tumor stage I.

**Table 4-2.** Results of statistical analysis of dependent variables for tumor stage II.

| Variables | Total<br>(n = 57) | ER group<br>(n = 30) | Non-ER<br>group<br>(n = 27) | p | statistic |
| --- | --- | --- | --- | --- | --- |
| Hospital<br>Costs_CNY,<br>Mean ± SD | 74503.05 ±<br>9259.14 | 75576.01 ±<br>10654.74 | 73310.87 ±<br>7431.45 | 0.361 | -0.921 |
| Length of Hospital<br>Stay_d,<br>Mean ± SD | 16.61 ± 3.73 | 16.47 ± 2.69 | 16.78 ± 4.68 | 0.756 | 0.312 |
| TFFFAS_d,<br>Mean ± SD | 4.57 ± 2.30 | 4.15 ± 1.05 | 5.05 ± 3.11 | 0.161 | 1.437 |
| TFSFAS_d,<br>Mean ± SD | 6.85 ± 2.59 | 6.40 ± 1.21 | 7.35 ± 3.51 | 0.191 | 1.335 |
| Postoperative<br>Complications, n (%) |  |  |  | 1.000 | Fisher's<br>exact test |
| NO = 0 | 53 (93.0) | 28 (93.3) | 25 (92.6) |  |  |
| YES = 1 | 4 (7.0) | 2 (6.7) | 2 (7.4) |  |  |

**Table 4-3.** Results of statistical analysis of dependent variables for tumor stage III.

| Variables | Total<br>(n = 54) | ER group<br>(n = 25) | Non-ER<br>group<br>(n = 29) | p | statistic |
| --- | --- | --- | --- | --- | --- |
| Hospital Costs_CNY,<br>Mean ± SD | 69929.01 ±<br>9049.86 | 71443.97 ±<br>9574.28 | 68623.01 ±<br>8522.77 | 0.257 | -1.146 |
| Length of Hospital<br>Stay_d, Mean ± SD | 15.96 ± 4.74 | 17.20 ±<br>5.86 | 14.90 ± 3.26 | 0.075 | -1.818 |
| TFFAS_d,<br>Mean ± SD | 4.52 ± 2.20 | 5.10 ± 2.52 | 4.01 ± 1.79 | 0.069 | -1.855 |
| TFSFAS_d,<br>Mean ± SD | 7.33 ± 3.43 | 8.35 ± 4.19 | 6.45 ± 2.33 | 0.041 | -2.096 |
| Postoperative<br>Complications, n (%) |  |  |  | 1.000 | Fisher's exact<br>test |
| NO = 0 | 49 (90.7) | 23 (92.0) | 26 (89.7) |  |  |
| YES = 1 | 5 (9.3) | 2 (8.0) | 3 (10.3) |  |  |

**Table 4-4.** Results of statistical analysis of the dependent variable for tumor stage as unspecified stage.

| Variables | Total<br>(n = 179) | ER group<br>(n = 92) | Non-ER<br>group<br>(n = 87) | p | statistic |
| --- | --- | --- | --- | --- | --- |
| Hospital<br>Costs_CNY,<br>Mean ± SD | 72578.51 ±<br>10294.67 | 75510.44 ±<br>11584.95 | 69478.08 ±<br>7643.31 | <0.001 | -4.133 |
| Length of Hospital<br>Stay_d,<br>Mean ± SD | 15.41 ± 3.694 | 16.29 ±<br>3.957 | 14.47 ± 3.154 | <0.001 | -3.416 |
| TFFFAS_d,<br>Mean ± SD | 3.98 ± 2.01 | 4.25 ± 2.11 | 3.69 ± 1.86 | 0.066 | -1.853 |
| TFSFAS_d,<br>Mean ± SD | 6.20 ± 2.04 | 6.51 ± 2.18 | 5.87 ± 1.85 | 0.034 | -2.131 |
| Postoperative<br>Complications,<br>n (%) |  |  |  | 0.768 | Fisher's exact<br>test |
| NO = 0 | 167 (93.3) | 85 (92.4) | 82 (94.3) |  |  |
| YES = 1 | 12 (6.7) | 7 (7.6) | 5 (5.7) |  |  |

## Discussion

ER is characterised by its pungent nature and spicy, bitter taste with low toxicity. From a traditional Chinese medicine review, it is associated with the liver, spleen, stomach, and kidney meridians. Its effects include dispersing cold and relieving pain, reducing rebellious qi and stopping vomiting, and aiding yang tonification and stopping diarrhoea[29]. The “Shennong Bencao Jing” also describes ER as a mildly warm herb, mainly used for warming the middle, dispelling cold, removing dampness and blood stasis, expelling wind evil and opening the body channels, and classifies it as a medium- grade herb[16].

The Shen Que acupoint of the Ren meridian is located at the navel. The Ren meridian, one of the eight extraordinary meridians, intersects with the twelve ordinary meridians and serves as the hub of the meridian system, connecting hundreds of vessels and penetrating the five zang organs and six fu organs[30–31]. Applying medicine to the navel can promote circulation in the meridians, harmonise abdominal qi and relieve abdominal bloating and discomfort[31].

Modern research has found that during embryonic development, the navel is the final site of closure of the abdominal wall, lacking subcutaneous fat but containing abundant blood vessels, lymphatic vessels and nerves, making it highly permeable[32–34]. Therefore, drug molecules can easily penetrate the epidermal keratin layer of the umbilicus, rapidly infiltrate the intercellular space and exert their effects. In clinical practice, the commonly used ER hot compress therapy uses local heat to facilitate the penetration of drugs such as evodiamine and rutaecarpine from the ER into the abdomen (around the navel) to accelerate the recovery of post-operative gastrointestinal function[35–36].

This retrospective cohort study aimed to investigate the effects of using ER hot compress therapy on the recovery of gastrointestinal (GI) function in patients undergoing colorectal cancer surgery. Currently, most retrospective cohort studies tend to directly analyse the raw data to verify the effectiveness of ER hot compress therapy. However, due to limitations in study design, those studies suffer from recall bias and multiple confounding factors that do not adequately account for the importance of ER hot compress therapy in the recovery of postoperative gastrointestinal function. To reduce these confounding factors, this study adopted the PSM method, which is commonly used in clinical practice, and investigated the factors that influence the efficacy of ER hot compresses. At the same time, a subgroup analysis according to tumor stage was performed to gain insight into the use of the method in patients with different stages of the disease.

The results of our study showed that when comparing patients in the ER and non-ER groups, the matched results showed that there was no significant difference between the two groups in terms of indicators related to gastrointestinal function after laparoscopic surgery (p-values were all greater than 0.05), which was similar to the results of the studies by Chen[37] and Jian[38]. Interestingly, however, when analysing the tumor stage into subgroups according to the AJCC TNM system[28], we found that the mean postoperative recovery time on a full-liquid diet was shorter in the ER group for patients with tumor stage II compared to the non-ER group (non-ER group: 5.05 ± 3.11 vs. ER group: 4.15 ± 1.05), the mean recovery time on semi-liquid diet was also shorter (non-ER group: 7.35 ± 3.51 vs. ER group: 6.40 ± 1.21), and the mean length of hospital stay was also shorter (non-ER group: 16.78 ± 4.68 vs. ER group: 16.47 ± 2.69). Although not statistically significant, these findings imply that patients with different tumor stages may differ in the therapeutic efficacy of ER hot compress therapy, suggesting that postoperative patients with early-stage tumors may be a potential population to benefit from ER hot compress therapy.

In addition, the combination of ER hot compress therapy with other complementary and alternative therapies has been widely used after a variety of abdominal surgeries[39–43]. These therapies include pain management[44], ultrasound electroconductive transdermal drug delivery techniques[45], moxibustion[46], auricular acupressure and electroacupuncture[47–48]. The results of clinical studies have shown that ER hot compress therapy combined with other therapies can not only improve the symptoms of urinary retention, nausea and vomiting, abdominal distension and abdominal pain in postoperative patients, but also promote the recovery of gastrointestinal function in postoperative patients. However, because more than two alternative therapies were combined, it was not possible to determine exactly which therapy had a significant effect. Furthermore, although previous studies have explored the impact of different treatment options on patient recovery, few studies have analysed the cost-effectiveness of patients receiving complementary and alternative therapies from an economic perspective, especially when looking at hospital costs and treatment outcomes.

Therefore, our study filled the gap in previous research by conducting an in-depth analysis of the cost-effectiveness of patients receiving complementary and alternative therapies from an economic perspective. With a focus on assessing the economic impact of these therapies, particular attention was paid to hospital costs, the length of hospital stay and treatment outcomes. Our results indicate that patients receiving ER hot compress therapy showed an overall increasing trend in hospital costs. However, further analysis showed that there were no significant differences in hospital costs and the length of hospital stay between the ER and non-ER groups in different tumor stage subgroups (including tumor stages I and II as well as stage III), with p-values all greater than 0.2. Additionally, the length of hospital stay was not statistically significant for patients with tumor stage I or II (p > 0.1). This finding suggests that although receiving ER hot compress therapy may slightly increase overall patient costs, there was no apparent economic disadvantage within specific subgroups. Therefore, our study highlights the considerations that healthcare practitioners need to address how to effectively use treatment modalities and techniques to improve patient recovery rates and reduce hospital length of stay without significantly increasing patient costs.

## Limitations

The limitations of this study may include: (1) due to inherent methodological limitations of retrospective study, the accuracy of recording time of first postoperative flatus, first postoperative defecation and recovery of postoperative bowel sound was limited, which may have led to bias in the assessment of recovery of postoperative gastrointestinal function, potentially affecting the reliability of the study conclusions. (2) there was no statistical analysis of the differences in surgical experience between surgeons, nor was there a detailed investigation of potential confounding factors such as age and gender. (3) some objective laboratory indicators to validate the efficacy of ER hot compress therapy were not included. (4) the frequency of use of ER hot compress therapy by patients was not discussed and the effect of frequency of use on efficacy remains unclear. In addition, there may have been variability in the implementation of ER hot compress therapy. (5) patients were not followed up, which might limit understanding of post-discharge outcomes. (6) selection bias could not be avoidedi.e. patients with more severe conditions are more likely to choose complementary and alternative therapies such as ER hot compress therapy. This may lead to selection bias and might affect the results of the trial.

Therefore, we recommend the following.

First, plarger sample sizes, multicentre controlled prospective cohort studies or randomised controlled trials are recommended. Second, future clinical studies on ER hot compress therapy should explicitly state that the treatment team’s characteristics and the experiences of the surgeon to better control for potential confounding factors. Third, the objective experimental indicators should be selected and not limited to comparing the recovery of gastrointestinal function during hospitalisation after surgery, follow-up observations, such as rehospitalisation rates and outpatient follow-up, should be included to complement the assessment of postoperative recovery of gastrointestinal function. Fourth, future studies should provide a more detailed record of the frequency of ER hot compress therapy usage and explore how usage frequency affects treatment efficacy. If possible, the overall effects on the recovery of postoperative gastrointestinal function of different groups should be compared by follow-up after discharge. Finally, future studies can employ qualitative research methods, such as interviewing patients about their experiences of postoperative use of ER hot compress therapy, to comprehensively evaluate the clinical benefits of this complementary and alternative therapy.

## Conclusion

The overall comparison suggests that Evodia rutaecarpa (ER) hot compress therapy may not significantly promote recovery of gastrointestinal function, however, patients with early-stage colorectal cancer may be a potential beneficiary group. Therefore, healthcare providers should consider treatment diversity and patient characteristics when formulating personalised treatment plans. By developing treatment regimes based on scientific evidence and individual patient needs, healthcare professionals can optimise treatment outcomes and effectively meet patient needs.

## Data Availability

All relevant data are within the manuscript and its Supporting Information files.

## Acknowledgments

We would like to thank Mr. XiaoFeng Chen for his technical support and guidance.

## Supporting information

S1 File. The Strengthening the Reporting of Observational Studies in Epidemiology (STROBE) guidelines.

S1 Fig. The density plot of the ER group and non-ER group before and after PSM.

S2 Fig. The eCDF plot of the ER group and non-ER group before and after PSM.

S3 Fig. Differences in standard deviation plots of the ER group and non-ER group before and after PSM under different caliper values.

S4 Fig. Histograms of the ER group and non-ER group before and after PSM at different caliper values.

S1 Table. The 95% confidence intervals for continuous variables before and after PSM.

S2 Table. Types of postoperative complications.

